# Prevalence and Patterns of Lumbar Spine Modic Changes Among Adult Patients with a History of Low Back Pain in the Northern Zone, Tanzania

**DOI:** 10.64898/2026.09.24.26363960

**Authors:** Joseph Lutatina, Rogers Temu, Peter M. Magembe, Faiton N. Mandari, Langas Majuka, Deogratius Mtui, Joshua Mollel, Frank Olottu, Felister Uisso, Godlisten Kawiche, Mathias S. Ncheye, Honest H. Massawe

## Abstract

**Introduction:** Low back pain (LBP) is a common leading cause of functional disability in adults worldwide, presenting with Modic changes that are visible within the endplates of the vertebral body under magnetic resonance imaging (MRI). Modic changes (MC) are not just a coincidental imaging finding; they reflect underlying pathology that needs treatment focus. This study aimed to ascertain the prevalence and patterns of Modic changes in the lumbar spine among adult patients with a history of low back pain attending the Kilimanjaro Christian Medical Centre (KCMC) Orthopedic clinic.

**Methods:** A descriptive cross-sectional study was conducted at the KCMC orthopedic clinic from 2^nd^ October 2024 to 30^th^ May 2025. Data from adult patients with a history of low back pain who had undergone lumbar spine MRI were collected, cleaned, and analyzed in SPSS (version 25).

Categorical variables were summarized by frequency and proportion, while continuous variables were summarized using the mean and standard deviation. Frequency tables, histograms, and pie charts were used for results presentation.

**Results:** Of the 283 patients enrolled in the study, the mean age was (57.2 ±15.7) years, with the majority (145; 51.2%) being aged > 59 years. Most patients, 203 (71.7%), were females, while 176 (62.2%) were married, and 142 (50.2%) were overweight. The prevalence of MC on the lumbar spine among patients with a history of low back pain was 66.4% (n=188). Among patients with lumbar spine MC, the majority, 112 (59.6%), were aged over 59 years, and 125 (66.5%) had lumbar spine MC type II, which was the most predominant compared to other types of MC.

**Conclusion:** Modic changes on the lumbar spine are prevalent in adult patients with low back pain, particularly those over 59 years, with type II being the most common. Healthcare professionals and policymakers should recognize Modic changes as clinically relevant findings that require consideration in the management of patients with a history of low back pain rather than dismissing them as mere incidental or coincidental observations.

## Introduction

All over the world, low back pain (LBP) is the leading cause of functional disability. In the coming decades, it is anticipated that the overall burden of LBP as well as the expenses associated with illness would rise even more [1]. Lumbar degenerative disease is one of the leading causes of low back pain. As MRI technology continues to advance, researchers have focused more on the connection between low back pain, endplates, and vertebrae. Modic changes are the specific bone marrow lesions visible within the endplates of the vertebral body on magnetic resonance imaging (MRI) [2]. Modic changes occur as a series of pathological alterations when the cartilage endplate loses its protective function, leading to inflammation in the nearby spongy bone, followed by fat infiltration in the vertebral body, which eventually leads to scarring and calcification. These progressive changes are categorized into three groups [3]. On T1-weighted imaging, type I Modic changes appear hypointense, while on T2-weighted imaging they appear hyperintense, due to acute inflammation or oedema within the vertebral endplate. Modic changes type II is characterized by hyperintense signals on both T1-weighted and T2 -weighted MRI scans, suggesting fatty degeneration or endplate necrosis in the bone marrow. Also, Modic changes type III are characterized by hypointense signals on T1-weighted and T2-weighted imaging, reflecting subchondral bone necrosis [4]. Globally, particularly in Asia, over 60% of individuals with chronic low back pain exhibit Modic changes [5]. In Africa, particularly in Nigeria, Modic changes were found in more than 23% of adults with chronic low back pain; however, type I Modic changes were the most prevalent [6]. Among individuals with a history of low back pain, Modic changes are probably not just a coincidental imaging finding; rather, they reflect an underlying pathology that ought to be a focus of treatment [7]. Information is scarce regarding the understanding and consideration of the relation between Modic changes on the lumbar spine and low back pain symptoms. This study aims to fill this knowledge gap by determining the prevalence and patterns of Modic changes on the lumbar spine among patients with a history of low back pain attending the KCMC Orthopedic Clinic from October 2024 to May 2025. Recognizing the prevalence and patterns of lumbar spine Modic changes among patients with a history of LBP will help to create awareness about Modic changes, which is much more important for developing the management guidelines and protocols used by doctors, physiotherapists, occupational therapists, and policymakers for better improvement of clinical decision-making when treating patients with low back pain.

## Material and Methods

### Study design

A hospital-based cross-sectional study was conducted to assess the prevalence and patterns of lumbar spine Modic changes among adult patients with a history of low back pain.

### Study setting and period

Data were collected from 2^nd^ October 2024 to 30^th^ May 2025 at KCMC, a zonal referral hospital located in Moshi Municipal, Northern Tanzania, in the Kilimanjaro region. It serves more than 11 million people, primarily serving residents of Kilimanjaro, Arusha, Tanga, Manyara, Singida, and Tabora. Currently, it receives around 1000 outpatients daily; its inpatient admission capacity for inpatients is 450 patients, although it is often stretched to accommodate 150-200 more. KCMC is the only referral hospital in Tanzania’s northern zone, which also receives patients from neighboring regions of Tanzania. Due to its geographical location bordering Kenya, KCMC also receives patients from across the border. KCMC Orthopedic Department was established in 1986, and comprises 60 inpatient beds distributed across six general and four private bays. However, the number of patients frequently exceeds the available bed capacity. KCMC Orthopedic Clinic can accommodate up to ten patients at once and is open for more than eight hours per day, five days a week.

### Study population

The study enrolled all adult patients (>18 years) attending KCMC Orthopedic Clinic who presented with a history of low back pain and who had done a lumbar spine MRI in the past three months during the study period. Patients with a history of spine injury, spine surgery and who were previously diagnosed with spine disorders such as a spine tumor or spine infection were excluded

### Sample size determination

The sample size was determined by taking a prevalence value of 23.8% (0.238) from a cross-sectional study conducted in North Central Nigeria, considering a confidence interval (CI) of 95% and a margin of error of 5% [6]. N was calculated using the Cochran formula

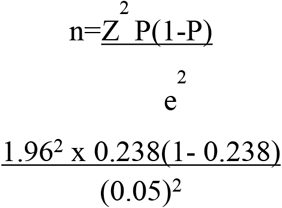

Where n = sample size; P = estimated prevalence, which was 23.8%; Z = level of confidence interval, which is equal to 1.96; e = margin of error or desired precision, which is equal to 5%=0.05, a total sample size was 279.

### Sampling procedure

A convenience sampling method was used to select study participants based on a history of low back pain among adult patients attending the KCMC Orthopedic clinic and who had undergone lumbar spine MRI in the past three months during the study period.

### Study variables

#### The independent variables were

social demographic data including age, gender, Body Mass Index (kg/m^2^) and Marital status.

**The dependent variable was lumbar spine Modic changes**, defined as the prevalence and patterns of lumbar spine Modic changes among all adult patients with a history of low back pain.

### Data collection

A triage guide for enrollment of study participants based on inclusion and exclusion criteria was formulated. A researcher gave a triage guide to a triage nurse, and then all patients with a history of low back pain attending the Orthopedic clinic were screened. Patients who met the triage guide criteria were invited to participate in this study. Data were collected using a structured questionnaire, which was composed of sociodemographic characteristics and radiological information describing patterns of lumbar spine Modic changes. Structured questionnaires were available in both English and Swahili languages to allow the study participants to express themselves in either language; they were translated and validated into Swahili using a pretesting and cognitive interviewing method.

A total of 361 patients with a history of low back pain were screened at the orthopedics clinic, of which 78 patients were excluded from the study (41 patients had a history of lumbar spine injury, 20 patients had a history of spine surgery, 10 had a history of spine infection, and 7 had a history of spine tumor). Two hundred eighty-three (283) patients who met the inclusion criteria were consented; the researcher and research assistants interviewed the patients face-to-face, using a structured questionnaire. Lumbar spine MRI of the consented patients was reviewed with the assistance of both a senior orthopedics specialist and a radiologist.

### Data Analysis

Data were coded, entered, processed, and analyzed using the SPSS program version 25. Categorical variables were summarized by frequency and proportion, while measures of central tendency (mean) with respective measures of dispersion were used for numerical variables. Frequency tables, histograms, and pie charts were used for results presentation.

### Ethical consideration

The ethical clearance was obtained from KCMC University Research and Ethics Review Committee (No. PG149 / 2024). Authorization to conduct the study was also secured from the head of the Department of Orthopedics and Traumatology at KCMC, as well as from KCMC administration. Study participants provided written informed consent to participate voluntarily. The authors declared that the information presented in this research article is original work that has not been presented elsewhere.

## Results

### Demographic Characteristics of the study participants

This study included a total of 283 study participants. The mean (SD) age of the study participants was 57.2 (15.5) years. However, 145 (51.2%) were aged > 59 years, 203 (71.7%) were females, 176 (62.2%) were married, and 142 (50.2%) were overweight, as shown in Table 1.

**Table 1:**
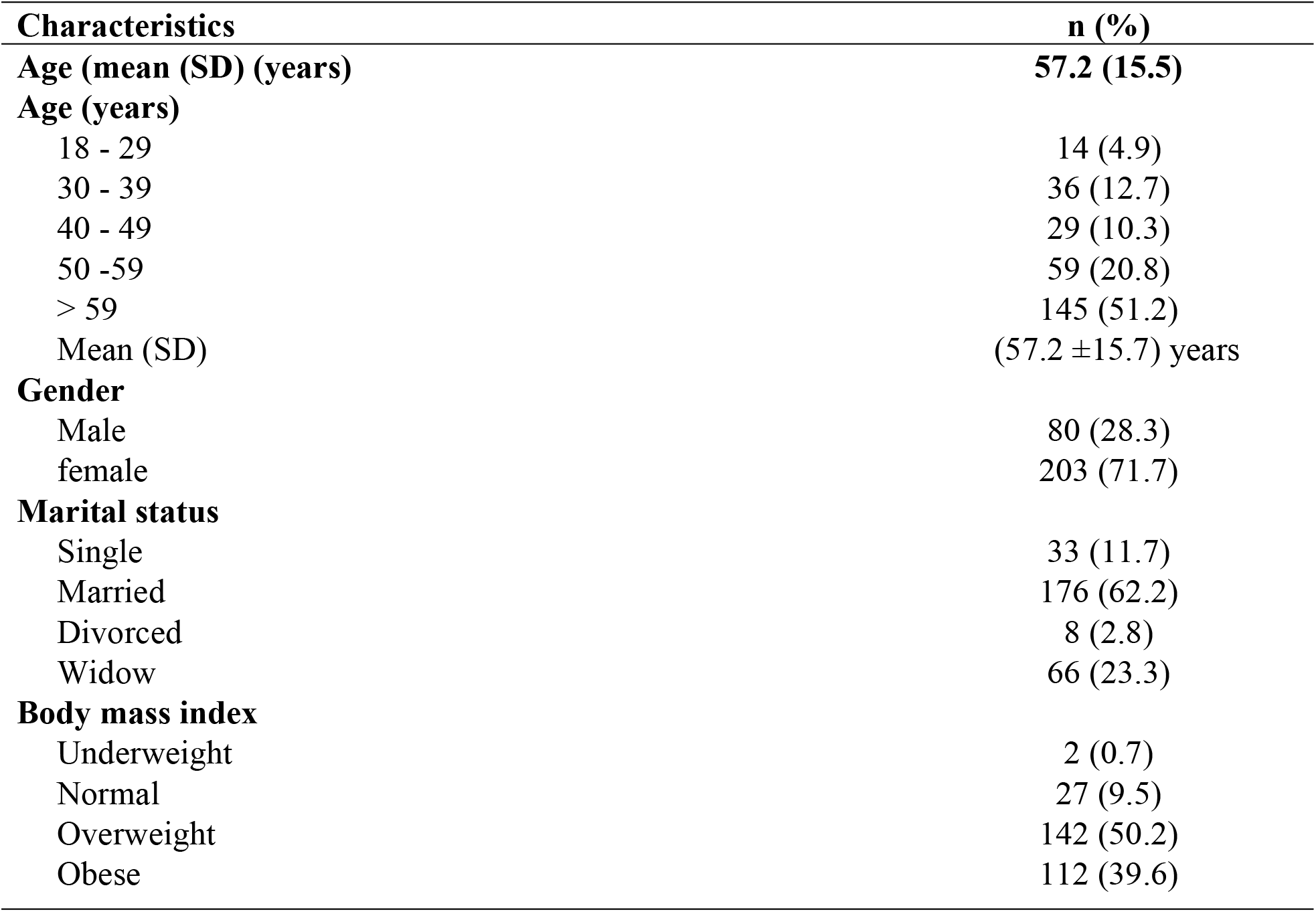
Characteristics of the study participants (n=283)

### The prevalence of Modic changes on the lumbar spine

The prevalence of Modic changes on the lumbar spine among patients with history of low back pain who attended Orthopaedic clinic from October 2024 to May 2025 was 66.4% (n=188), as illustrated in figure 1.

**Figure 1:**
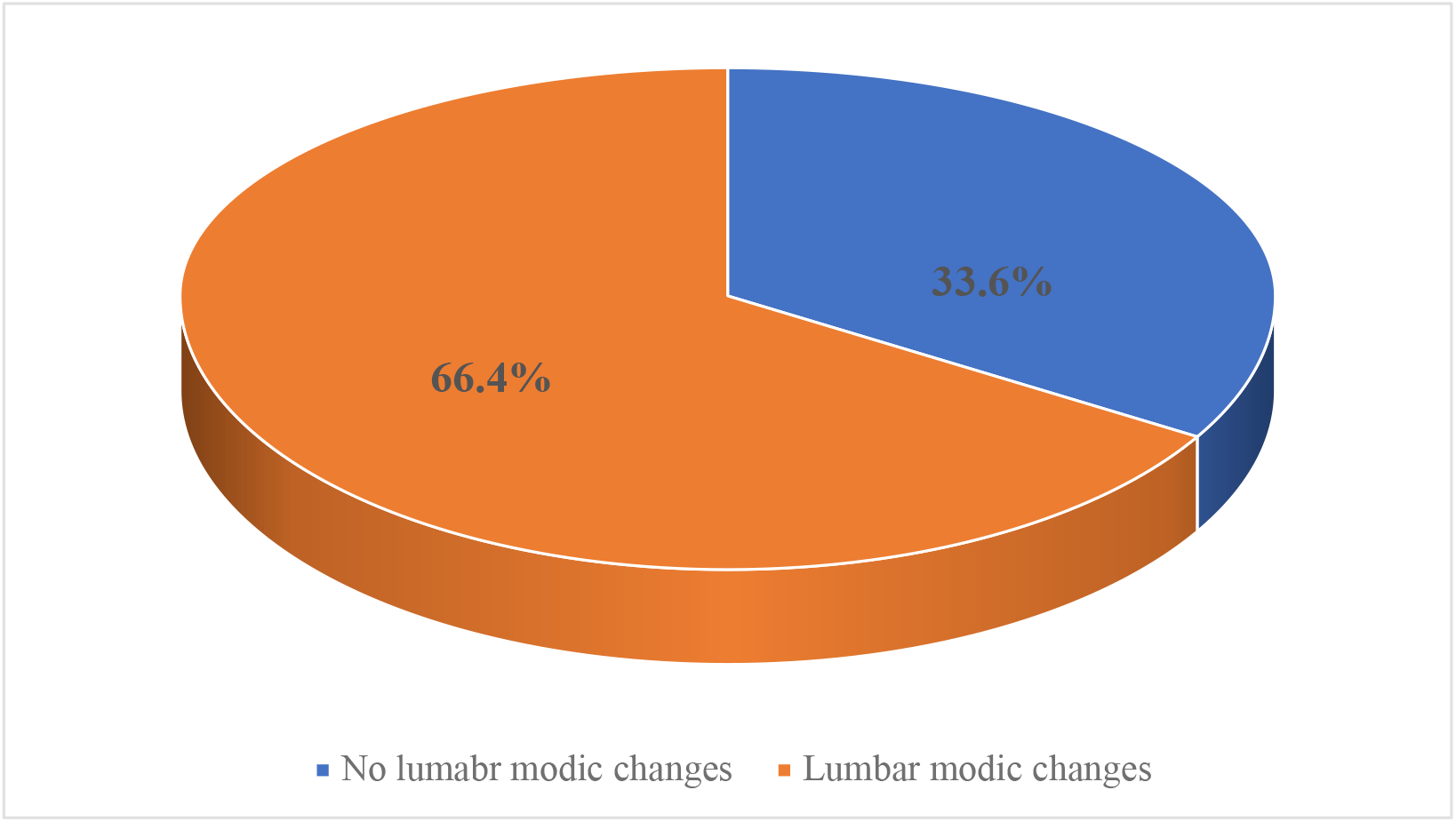
Prevalence of lumbar spine Modic changes (n=283)

**Figure 2:**
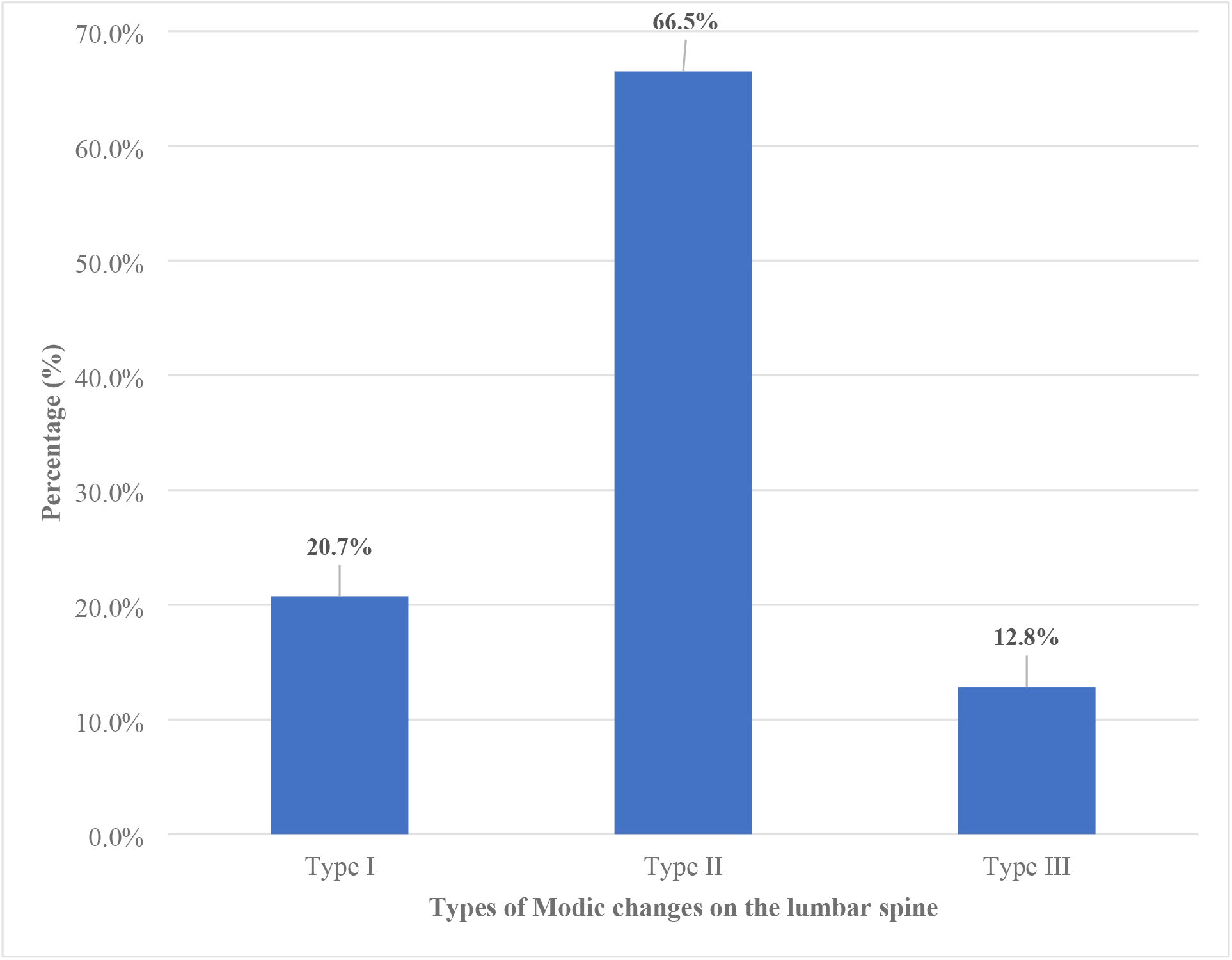
Types of lumbar spine Modic changes (n=188)

### Characteristics of the study participants with Modic changes on the lumbar spine

Among patients with lumbar spine Modic changes, 112 (59.6%) were aged over 59 years, 133 (70.7%) were female, 122 (64.9%) were married, and 92 (48.9%) were overweight, as shown in Table 2.

**Table 2:**
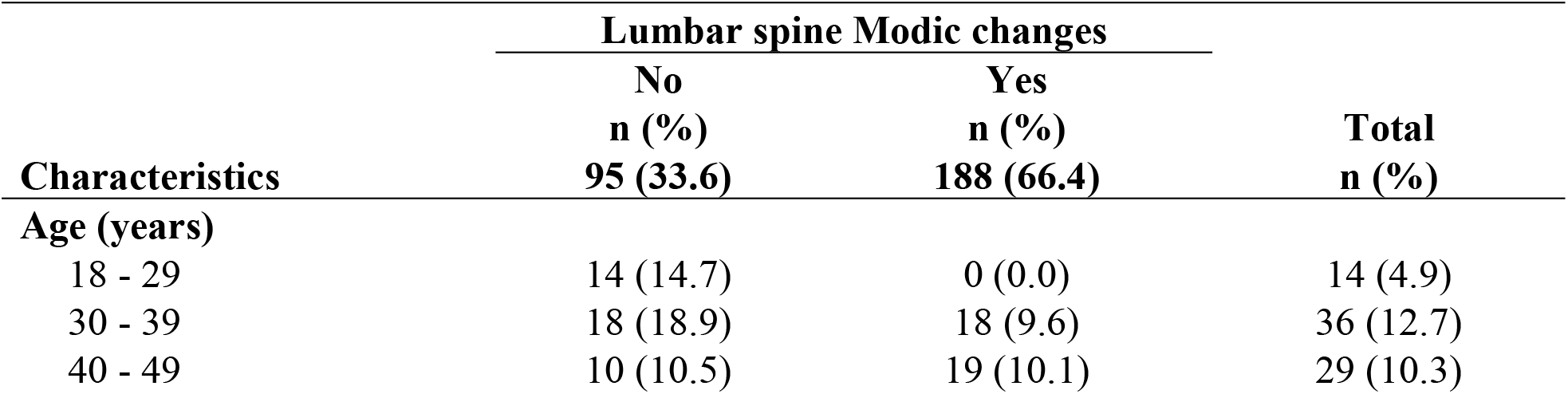
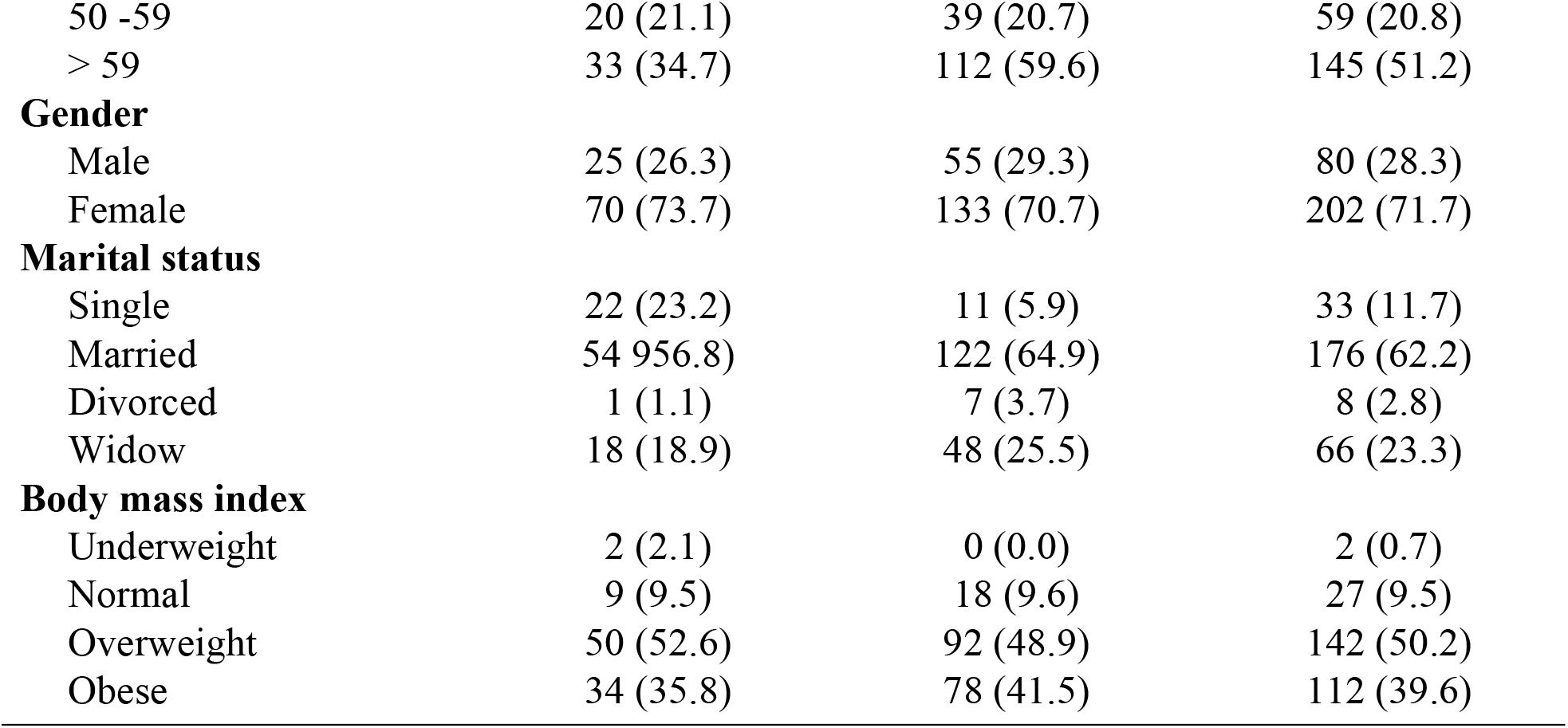
Characteristics of the study participants among those with lumbar Modic changes (n=188)

| Characteristics | Lumbar spine Modic changes |  | Total<br>n (%) |
| --- | --- | --- | --- |
|  | No<br>n (%) | Yes<br>n (%) |  |
|  | 95 (33.6) | 188 (66.4) |  |
| Age (years) |  |  |  |
| 18 - 29 | 14 (14.7) | 0 (0.0) | 14 (4.9) |
| 30 - 39 | 18 (18.9) | 18 (9.6) | 36 (12.7) |
| 40 - 49 | 10 (10.5) | 19 (10.1) | 29 (10.3) |
| 50 -59 | 20 (21.1) | 39 (20.7) | 59 (20.8) |
| > 59 | 33 (34.7) | 112 (59.6) | 145 (51.2) |
| **Gender** |  |  |  |
| Male | 25 (26.3) | 55 (29.3) | 80 (28.3) |
| Female | 70 (73.7) | 133 (70.7) | 202 (71.7) |
| **Marital status** |  |  |  |
| Single | 22 (23.2) | 11 (5.9) | 33 (11.7) |
| Married | 54 (56.8) | 122 (64.9) | 176 (62.2) |
| Divorced | 1 (1.1) | 7 (3.7) | 8 (2.8) |
| Widow | 18 (18.9) | 48 (25.5) | 66 (23.3) |
| **Body mass index** |  |  |  |
| Underweight | 2 (2.1) | 0 (0.0) | 2 (0.7) |
| Normal | 9 (9.5) | 18 (9.6) | 27 (9.5) |
| Overweight | 50 (52.6) | 92 (48.9) | 142 (50.2) |
| Obese | 34 (35.8) | 78 (41.5) | 112 (39.6) |

### Types of Modic changes on the lumbar spine

Regarding types of lumbar Modic changes, 39 (20.7%) patients had lumbar spine Modic changes type I, 125 (66.5%) patients had lumbar spine Modic changes type II, and 24 (12.8%) patients had lumbar spine Modic changes type III, as illustrated in Figure 3.

**Figure 3:**
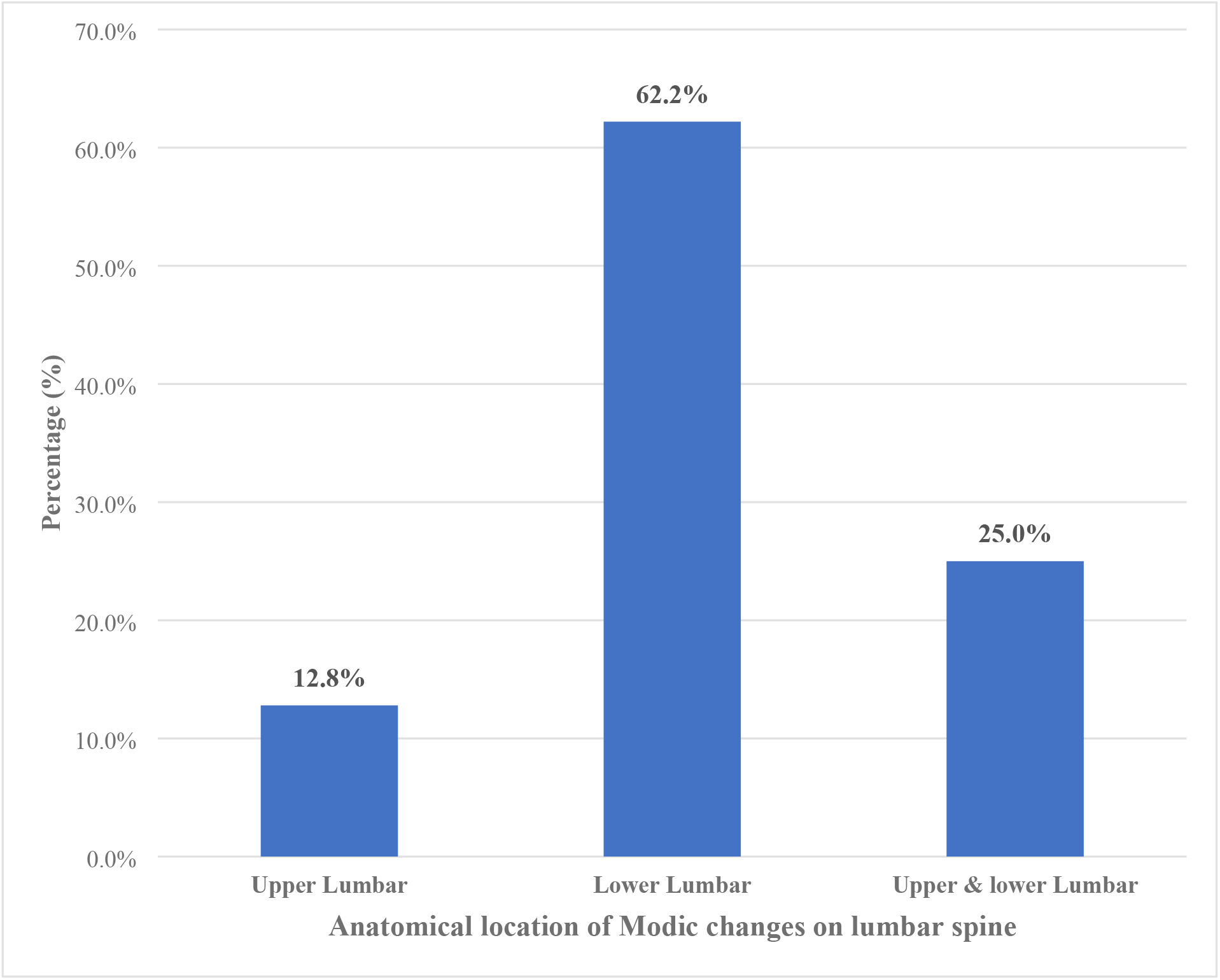
Anatomical location of Modic changes on the lumbar spine (n=188).

### Anatomical location of Modic changes on the lumbar spine

Regarding the anatomical location of Modic changes on the lumbar spine, 24 (12.8%) patients had Modic changes in the upper lumbar region, 117 (62.2%) had Modic changes in the lower lumbar region, and 47 (25%) had Modic changes in both the upper and lower lumbar regions as illustrated in Figure 4.

## 4. Discussion

This study aimed to assess the prevalence and patterns of lumbar spine Modic changes among adult patients with a history of low back pain attending KCMC Orthopedic Clinic. This study found that 66.4% (188 out of 283 patients) of patients with a history of low back pain had Modic changes in their lumbar spine I, which is higher and very significant, and suggests that Modic changes are highly prevalent. Although the prevalence is higher, it appears to be closer to the prevalence of 63.5% reported by [8] in a study that was conducted in Japan. This similarity was likely due to the use of the same MRI scanner, a 1.5 Tesla high-field scanner, which is a well-established imaging technology for identifying the presence and appearance of Modic changes. However, differences in prevalence were observed in three studies. One study conducted in Central Nigeria by [6] reported a prevalence of 23% (35 out of 147 patients); additionally, studies conducted in Finland by [9] and [10] revealed prevalences of 55.6% and 56%, respectively. This discrepancy in prevalence was probably due to factors such as the mean age and gender of the study participants, whereby the study done by [6] involved 147 patients with a mean age of 53.6. Also, a study done by [11] involved white men with a mean age of 47, and that of [12] involved 561 white men with a mean age of 49.8, while our current study involved 283 participants regardless of their gender with a mean age of 57.2. This highlights the clinical relevance and consideration of Modic changes when treating patients with a history of low back pain.

Also, in this study, 188 study participants had lumbar spine Modic changes, with 39 (20.7%) exhibiting type I, 125 (66.5%) exhibiting type II, and 24 (12.8%) exhibiting type III. Type II Modic changes have shown the highest prevalence compared to Modic changes type I and III, which may explain the chronicity of the disease. These findings were closed aligned to the findings reported by different studies such as studies conducted in Finland by [12] and [13], study done in Japan by [8] and another study done in China by [14]. All these studies found type II MC to be the most common type among the others. This similarity was likely due to the use of the same 1.5 Tesla high-field MRI scanner, which is highly effective and has a good capacity for identifying and classifying various forms of modic changes. A study done in Central Nigeria by [6] revealed a higher prevalence of type I modic changes, which was more than 39%, while type II and III modic changes had a prevalence of 30% each. This discrepancy might be due to the different MRI technology employed in these two studies. For example, a 0.2 Tesla low-field MRI scanner, which has limited capacity for identifying and classifying different types of modic changes, was employed in a study conducted in North Central Nigeria. In contrast, a 1.5 Tesla high-field MRI scanner was employed in the current study, utilizing advanced technology to identify and classify types of Modic changes, particularly in distinguishing type II modic changes. This highlights the relevance of using an advanced MRI scanner in identifying types of modic changes for better diagnosis and an appropriate management plan when treating patients with a history of low back pain.

In this study, the anatomical location of Modic changes on the lumbar spine was reported as follows: individuals with Modic changes on the upper lumbar region were 24 (12.8%), 117 (62.2%) had Modic changes on the lower lumbar region and 47 (25%) had modic changes on both the upper and lower lumbar region. These results indicate that most individuals exhibited Modic changes in the lower lumbar region, as this area is more susceptible to mechanical stress (axial loading). These findings were similar to the findings reported by [12] who reported that 74.6% had Modic changes in the lower lumbar region, but also, from another two studies conducted in China, one study was done by [15] reported that more than 75% of Modic changes were distributed to the lower lumbar region, and another study done by [14] reported that L4-L5 had 58 cases and L5-S1 had 72 cases, all of which correspond to more than 70% of the Modic changes in the lower lumbar region. These findings were similar to our study, likely because the lower lumbar region is a weight-bearing area that is consistently exposed to a higher percentage of biomechanical stress, which contributes to the degenerative process. This highlights that the lower lumbar segment is the weight-bearing part of the axial skeleton; therefore, much consideration should be taken when treating patients with a history of low back pain, for example, counselled patients about lifestyle modification, like restriction from hyperloading or lifting heavy weights, which will decelerate the degenerative process by decreasing mechanical stress to the lower lumbar region.

### Strength and limitation

The strength of this study included sample size obtained and used in this study was more than the estimated sample size, and all lumbar images were taken using a 1.5 Tesla high-field MRI, which is more advanced in identifying and classifying types of Modic changes.

Despite its strengths, this study has some limitations. This study employed a cross-sectional design, which prevented us from determining the causal relationship between lumbar spine Modic changes and low back pain. Additionally, this study constrained our capacity to assess the association between different Modic change patterns of the lumbar spine and variations in low back intensity and functional disability.

## Conclusion and recommendation

In this cross-sectional study, a significant prevalence of lumbar spine Modic changes was found, with Modic change type II being the most common. Healthcare professionals and policymakers should recognize Modic changes as clinically relevant findings that require consideration, appropriate evaluation, and management rather than dismissing them as mere incidental or coincidental observations. Another study should be done to analyze the association between patterns of lumbar spine Modic changes with both low back pain intensity and functional disability among adult patients with a history of low back pain. Also, the 1.5 T high-field MRI scanner used in this study has brought significant relevance in identifying and classifying types of Modic changes, which is more significant in clinical diagnosis and management planning. Therefore, we recommend the deployment of this type of MRI scanner in all other referral hospitals in Tanzania.

## Data Availability

The data used to generate this study have been presented within the manuscript

## Acknowledgements

We express our heartfelt thanks to all individuals who participated in the study: respondents, data collectors, and administrative officials.

## List of observations

DDD: Degenerative disc disease
KCMC: Kilimanjaro Christian Medical Centre
LBP: Low back pain
MC: Modic Changes
MRI: Magnetic Resonance Imaging
SPSS: Statistical Package for the Social Sciences
WHO: World Health Organisation

## Authors’ Contributions

**Conceptualization:** Joseph Lutatina, Honest H. Massawe and Peter M. Magembe

**Data curation:** Frank Olottu, Deogratius Mtui

**Formal analysis:** Joshua Mollel, Langas Majuka

**Investigation:** Felister Uisso, Peter M. Magembe, Mathias S. Ncheye and Honest H. Massawe

**Methodology:** Joseph Lutatina, Frank Olottu, Joshua Mollel

**Project administration:** Honest H. Massawe and Faiton Mandari

**Resources:** Honest H. Massawe and Faiton N. Mandari

**Software:** Joseph Lutatina, Frank Olottu, Joshua Mollel and Godlisten Kawiche

**Supervision:** Honest H. Massawe, Rogers Temu, Peter M. Magembe and Godlisten Kawiche

**Validation:** Joseph Lutatina, Mathias S. Ncheye and Faiton N. Mandari

**Writing original draft:** Joseph Lutatina and Peter M. Magembe

**Writing review and editing:** Joseph Lutatina, Honest H. Massawe, Rogers Temu and Peter M. Magembe

## Notes

### Competing Interest Statement

The authors have declared no competing interest.

